# Radiomics-Based Carotid Ultrasound Identifies Patterns Associated with Severe Coronary Calcification in the Absence of Carotid Plaque

**DOI:** 10.1101/2025.10.01.25337136

**Authors:** Youngjun Lee, Miran Lee, Rachel K. Surowiec, Jeonggyu Kang

**Affiliations:** Weldon School of Biomedical Engineering, Purdue University, West Lafayette, Indiana, United States; Total Healthcare Center, Kangbuk Samsung Hospital, Sungkyunkwan University School of Medicine, Seoul, Republic of Korea; Department of Electronic Engineering, Sogang University, Seoul, Republic of Korea; Department of Clinical Research Design & Evaluation, SAIHST, Sungkyunkwan University, Seoul, Republic of Korea

**Keywords:** Coronary Artery Calcium Score, Common Carotid Artery, Carotid Ultrasound, Radiomics, Feature Extraction, Feature Selection

## Abstract

**Background:** To assess whether radiomics analysis of carotid ultrasound (CUS) can identify texture features associated with severe coronary artery calcification, even in individuals without carotid plaque.

**Methods:** This study included 105 participants with coronary artery calcium score (CACS) > 400 and no carotid plaque, matched by age and sex to 105 controls with CACS=0. B-mode CUS images of the bilateral distal common carotid arteries (CCA) were analyzed, with 1-cm longitudinal regions of interest extending from the lumen to the adventitia. Radiomic features were extracted from each frame, filtered by variance and correlation, and ranked using bootstrap-based XGBoost feature importance (FI) and evaluated on internal and external datasets.

**Results:** Among 700 extracted features, the final retained features were reproducible: 8 (right) and 11 (left) for CACS=0, and 7 (right) and 10 (left) for CACS > 400 (all p < 0.05). Group-specific features, observed only in the CACS 0 or CACS > 400 group, included 90th Percentile (CACS=0: right distal CCA, FI=0.030; left distal CCA, FI=0.025) and Run Entropy (CACS > 400: right distal CCA, FI=0.046; left distal CCA, FI=0.038). Shared features such as Long Run Emphasis, Dependence Non-Uniformity, and Entropy were consistently observed across both groups and sides (FI=0.023-0.029), with Dependence Non-Uniformity consistent in the left distal CCA across both groups and datasets.

**Conclusions:** Although no plaque was detected on CUS, radiomics can identify ultrasound texture patterns associated with severe coronary calcification. This approach may improve detection of high-risk individuals who would otherwise be classified as low-risk by CUS alone.

**Graphic Abstract:** Texture-Based Radiomics of Carotid Ultrasound Reveals Severe Coronary Calcification. (A), Imaging modalities and study population: Adults undergoing health screenings at tertiary hospitals in City A and City B (2018–2022) who received same-day carotid ultrasound and CACS CT. After excluding plaque, CVD history, and poor image quality, 105 plaque-free participants with CACS > 400 were identified and age- and sex-matched to 105 plaque-free participants with CACS = 0. (B), Radiomics pipeline: carotid ultrasound images were segmented, preprocessed, filtered, and processed for feature extraction and selection, yielding reproducible texture features. (C), Radiomics feature maps: representative examples of plaque-free carotid ultrasound in patients with CACS = 0 and CACS >400. Entropy and dependence non-uniformity maps demonstrate distinct texture patterns associated with severe coronary calcification.

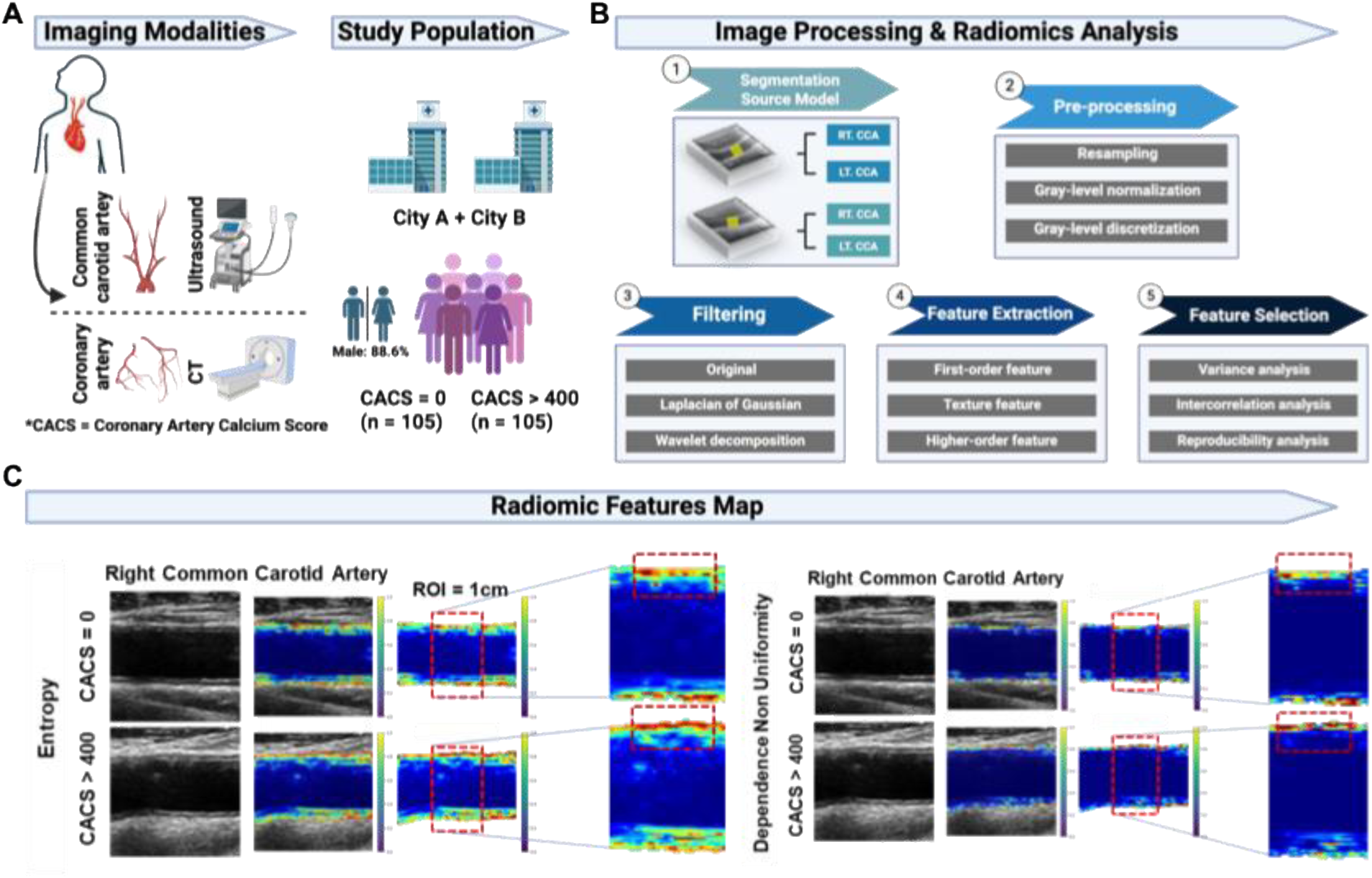

## INTRODUCTION

Cardiovascular disease (CVD) remains the leading cause of global mortality, with substantial events preventable through early risk stratification (1, 2). Coronary artery calcium score (CACS) through computed tomography (CT) and carotid plaque from carotid ultrasound (CUS) are validated markers of subclinical atherosclerosis that improve cardiovascular risk prediction beyond clinical risk scores alone (3–9). Although they reflect atherosclerotic burden in different vascular territories, CACS and carotid plaque offer complementary insights into systemic atherosclerosis.

CACS is guideline-endorsed for risk stratification but requires CT, which entails radiation exposure, higher cost, and limited accessibility in primary care (10–12). In contrast, CUS is portable, radiation-free, and cost-effective, enabling broader implementation in primary care and health screening settings (13, 14).

Although carotid plaque burden and CACS typically correlate, discordant findings are common (15, 16). In the BioImage study, 10.4% of participants showed positive CACS despite no detectable carotid plaque, whereas 19.6% had carotid plaque with CAC score of 0 (17). Notably, individuals without carotid plaque but with high CACS (upper tertile for age and sex) had nearly a ninefold higher incidence of major adverse cardiovascular events over three years compared with those with CACS = 0 (17). This discordance suggests that even without visible carotid plaque, carotid imaging may harbor subtle features reflecting severe coronary atherosclerosis, warranting investigation beyond conventional plaque assessment.

Radiomics is an emerging field that converts medical images into quantitative data by extracting numerous mathematical features related to image texture, shape, and intensity patterns (18). This approach can capture subtle imaging characteristics beyond traditional visual interpretation, potentially revealing subclinical atherosclerotic changes not apparent on standard imaging assessment and enabling more precise tissue characterization. In line with this potential, several studies have applied radiomics to cardiovascular imaging—primarily CT and MRI—to assess atherosclerotic plaque composition, coronary artery calcium, and disease severity, often improving risk prediction beyond conventional measures (19, 20). However, the application of radiomics to CUS, particularly for assessing associations with coronary atherosclerosis, has not been previously explored. This study aimed to determine whether radiomic features extracted from bilateral distal CCAs ultrasound cine images can differentiate patients with a CACS of 0 from those with a CACS > 400, even when no carotid plaque is detected on CUS, potentially addressing the underestimation of cardiovascular risk in individuals who are visually classified as low-risk via CUS alone.

## METHODS

This retrospective study was approved by the institutional review board (approval no. 2024-03-023); informed consent was waived due to the use of pre-existing, de-identified health screening data.

### 2.1. Study design and inclusion criteria

As shown in **Graphic Abstract A**, Data were from an ongoing cohort of adults (≥ 18 years) undergoing comprehensive health screenings at a tertiary referral hospital in City A and City B. Between January 2018 and December 2022, 29,579 participants underwent both CUS and CACS CT on the same day. Carotid plaque, defined as intima-media thickness (IMT) ≥ 1.5 mm according to the Mannheim consensus (21), was present in 12,126 participants, leaving 17,453 who were plaque-free. Among these, 123 had CACS > 400; after excluding those with CVD history (n=12) and poor image quality (n=6), 105 remained. From the plaque-free participants with CACS = 0 (n=13,735), 105 age- and sex-matched controls were selected (**Figure 1**). External validation included 272 patients from the same tertiary referral hospital who underwent CUS and either non-contrast coronary calcium CT or contrast-enhanced CT angiography within 90 days; 6 plaque-free with CACS > 400 and 9 plaque-free with CACS = 0 were included. Details of blood pressure measurement and biochemical analysis protocols are provided in the **Supplemental 1.1.**

**Figure 1.**
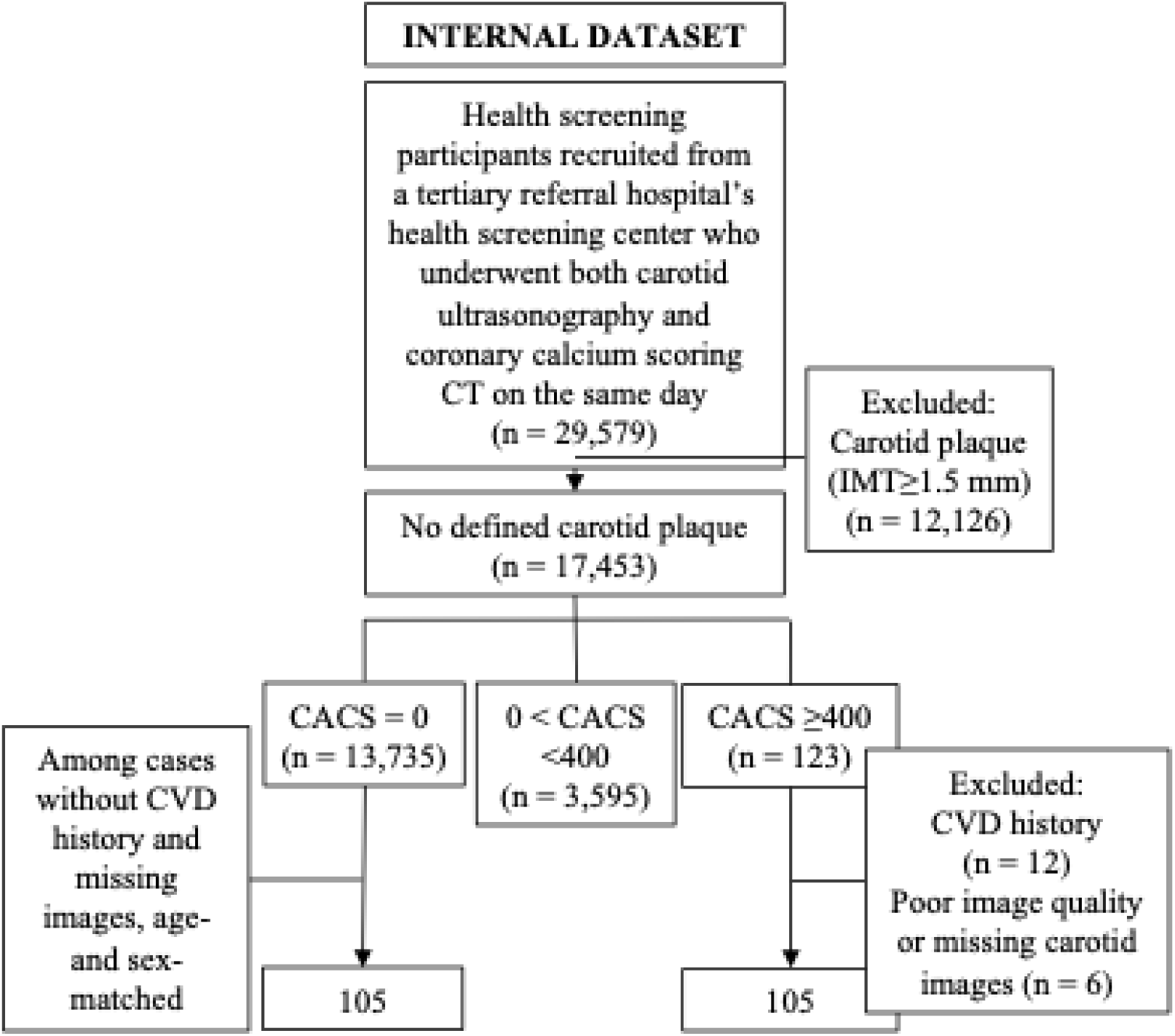
Flow diagrams of the study population selection. Internal dataset: Health screening participants (n = 29,579) from a tertiary referral hospital in City A and City B (2018–2022) who underwent both CUS and CACS CT on the same day. After excluding individuals with carotid plaque, CVD history, or missing/poor-quality images, 105 participants with CACS = 0 and 105 with CACS > 400 were selected through age- and sex-matching (A). External dataset: Inpatients and outpatients (n = 275) from the same tertiary referral hospital (2018–2021) who underwent both tests within a 90-day interval. After applying the same exclusion criteria, 9 participants with CACS = 0 and 6 with CACS > 400 were included (B). Abbreviation: CACS, coronary artery calcium score; CUS; CVD,

### 2.2 CACS measurements and CUS

CACS was quantified using the Agatston method on pre-contrast images from either non-contrast CT or CT angiography (22). The internal dataset used non-contrast cardiac CT, while the external dataset used pre-contrast images from contrast-enhanced coronary CT angiography. CUS was performed bilaterally using high-resolution B-mode scanning according to Mannheim consensus criteria for plaque definition, with far-wall mean IMT, peak systolic velocity (PSV), and end-diastolic velocity (EDV) measured at the distal CCA. Details on imaging equipment, acquisition parameters, and reader qualifications are provided in **Supplemental 1.2.**

### 2.3. Automated segmentation for region of interest (ROI) definition

To ensure accurate radiomics feature extraction, an automated segmentation method was developed to define the ROI, extending from the carotid artery lumen to the outer boundary of the adventitia. The ROI was standardized as a 1 cm segment proximal to a reference point located 1 cm distal to the bulb–CCA junction, identified by a qualified diagnostic cardiac sonographer. A U-Net architecture with a ResNet-34 encoder was used for segmentation, with performance evaluated using both internal and external validation datasets. Detailed acquisition parameters, dataset characteristics, and evaluation metrics are provided in Supplemental 1.3.

### 2.4. Radiomic feature extraction

For preprocessing, images were resampled when applicable to ensure consistent spatial resolution, following current recommendations (23). Pixel intensities were standardized using z-score normalization, scaling values between 0 and 1. To determine an appropriate bin width for gray-level discretization, we analyzed the intensity range in the training set only, thereby avoiding information leakage from the test data. Candidate bin widths of 0.2, 0.3, 0.4, and 0.5 were evaluated. Feature extraction was performed on both the original and filtered images, including those processed with Laplacian of Gaussian filters (*σ* = 1–5) and wavelet transforms. All first-order and texture-based features, including second- and higher-order statistics, were extracted according to the PyRadiomics v3.1.0 guidelines, yielding 700 features initially per carotid side (24).

### 2.5. Feature selection and reproducibility filtering

A multi-step filtering process was applied independently for each carotid side (right and left). Feature filtering was performed using empirically defined threshold ranges (e.g., variance < 0.1–0.5; Pearson correlation > 0.5–0.9) through repeated iterations to eliminate redundant and low-information features (25). To enhance reproducibility, feature stability was assessed via bootstrap-based ranking, where FI scores were computed using an XGBoost classifier in each iteration (26). Features that consistently achieved high rankings across bootstrap samples were retained for subsequent statistical analysis, defined as those appearing in > 80% of bootstrap iterations and ranking within the top 10 features by mean feature importance (FI) for each carotid side.

### 2.6. Statistical analysis

Participant demographics and clinical characteristics were presented as mean ± standard deviation and compared between groups using the same statistical approach. Continuous variables were summarized as medians with interquartile ranges (1st–3rd quartiles), and categorical variables as counts with corresponding percentages. Data preparation and processing were performed using the pandas library in Python (27). Statistical comparisons of radiomic features and ultrasound-derived conventional measures-such as IMT, PSV, and EDV-were conducted independently for each carotid side between the CACS = 0 and CACS > 400 groups. Normality was assessed using the Shapiro-Wilk test (*p* > 0.05 indicating normal distribution). For normally distributed variables, two-sample t-tests were applied with effect sizes calculated using Cohen’s *d*. For non-normally distributed variables, the Mann– Whitney U test was used, and effect sizes were reported as rank-biserial correlation (*r*) and Cliff’s delta (28). To visualize group differences, box plots of the top-ranked radiomic features were generated. All statistical analyses were performed using SPSS (version 28), with statistical significance set at *p* < 0.05.

## RESULTS

### 3.1. Participant Characteristics

The internal dataset included 210 individuals, consisting of 105 with CACS > 400 and 105 age- and sex-matched individuals with CACS = 0. The groups were age- and sex-matched, with a mean age of 58.1 ± 8.0 years and 88.6% men representation in both groups (*p* = 1.000 for both). No significant differences were observed in serum creatinine, cholesterol profiles, or blood urea nitrogen. However, the CACS > 400 group had significantly higher glycated hemoglobin (6.24 ± 1.17% vs. 5.65 ± 0.58%, *p* < 0.001) and systolic blood pressure (118.3 ± 11.9 mmHg vs. 113.9 ± 10.7 mmHg, *p* = 0.005).

In the external validation dataset, the mean age was higher in the CACS > 400 group (63.7 ± 6.3 years) than in the CACS = 0 group (54.1 ± 11.1 years), but this difference was not statistically significant (p = 0.103). No females were present in the CACS > 400 group compared with 44.4% in the CACS = 0 group (p = 0.018). Blood pressure was unavailable for this external dataset due to institutional constraints. (**Table 1**).

**Table 1.**
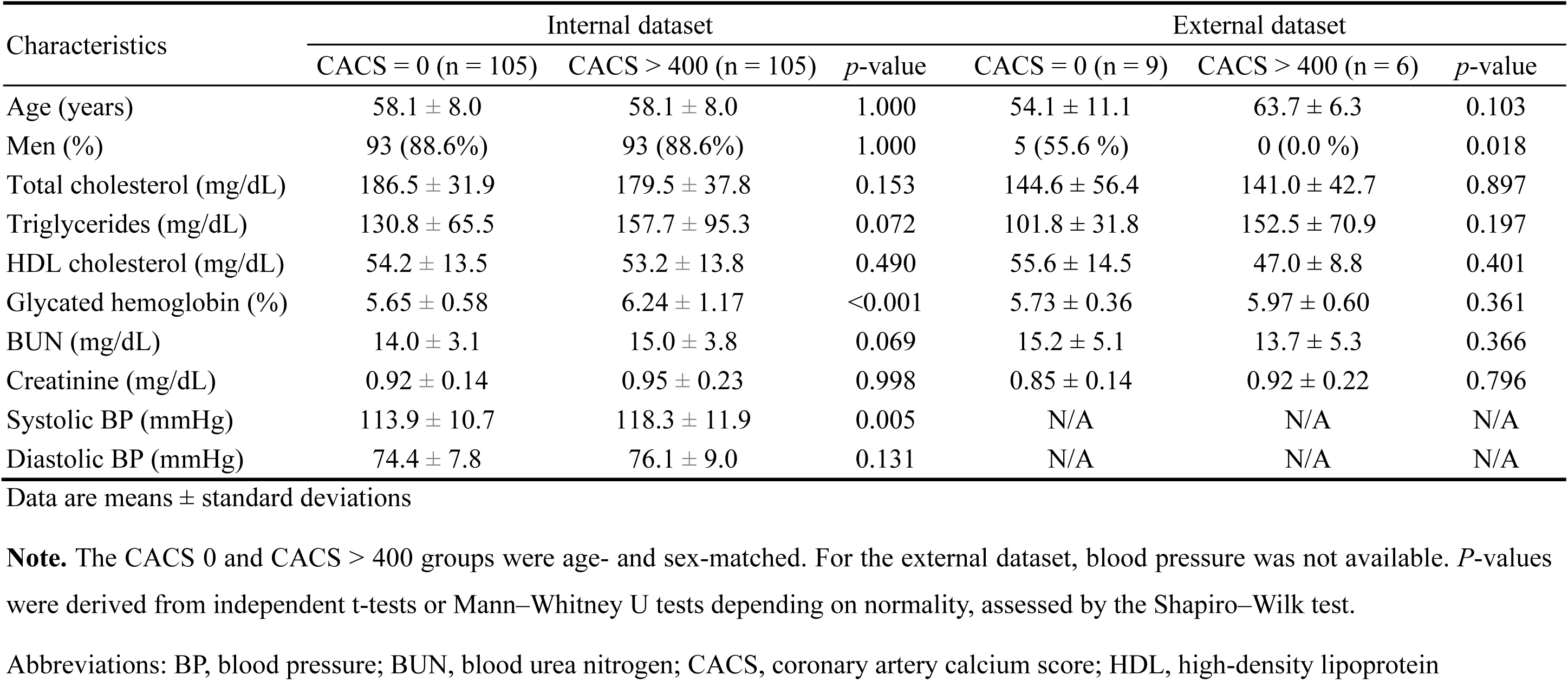
Participant characteristics.

### 3.2. Conventional CUS-derived measures

In the internal dataset (**Table 2**), statistically significant differences were observed in EDV between the CACS = 0 and CACS > 400 groups. Specifically, EDV was significantly lower in the CACS > 400 group in both the left distal CCA (21.7 ± 6.3 cm/s vs. 23.8 ± 6.6 cm/s, *p* = 0.032, *r* = 0.148) and the right distal CCA (20.7 ± 6.2 cm/s vs. 22.6 ± 5.4 cm/s, *p* = 0.008, *r* = 0.184). Other ultrasound-derived parameters, including IMT and PSV from both carotid arteries, did not show statistically significant differences (all *p* > 0.05), and corresponding effect sizes were small (*r* = 0.02 to 0.18). In the external dataset (**Table 2**), no statistically significant differences were found in IMT, PSV, or EDV between the two groups (all *p* > 0.05). Although EDV from the right distal CCA tended to be lower in the CACS > 400 group (13.7 ± 6.7 cm/s vs. 21.1 ± 9.7 cm/s, *p* = 0.091, Cohen’s *d* = 0.897), this difference did not reach statistical significance.

**Table 2.**
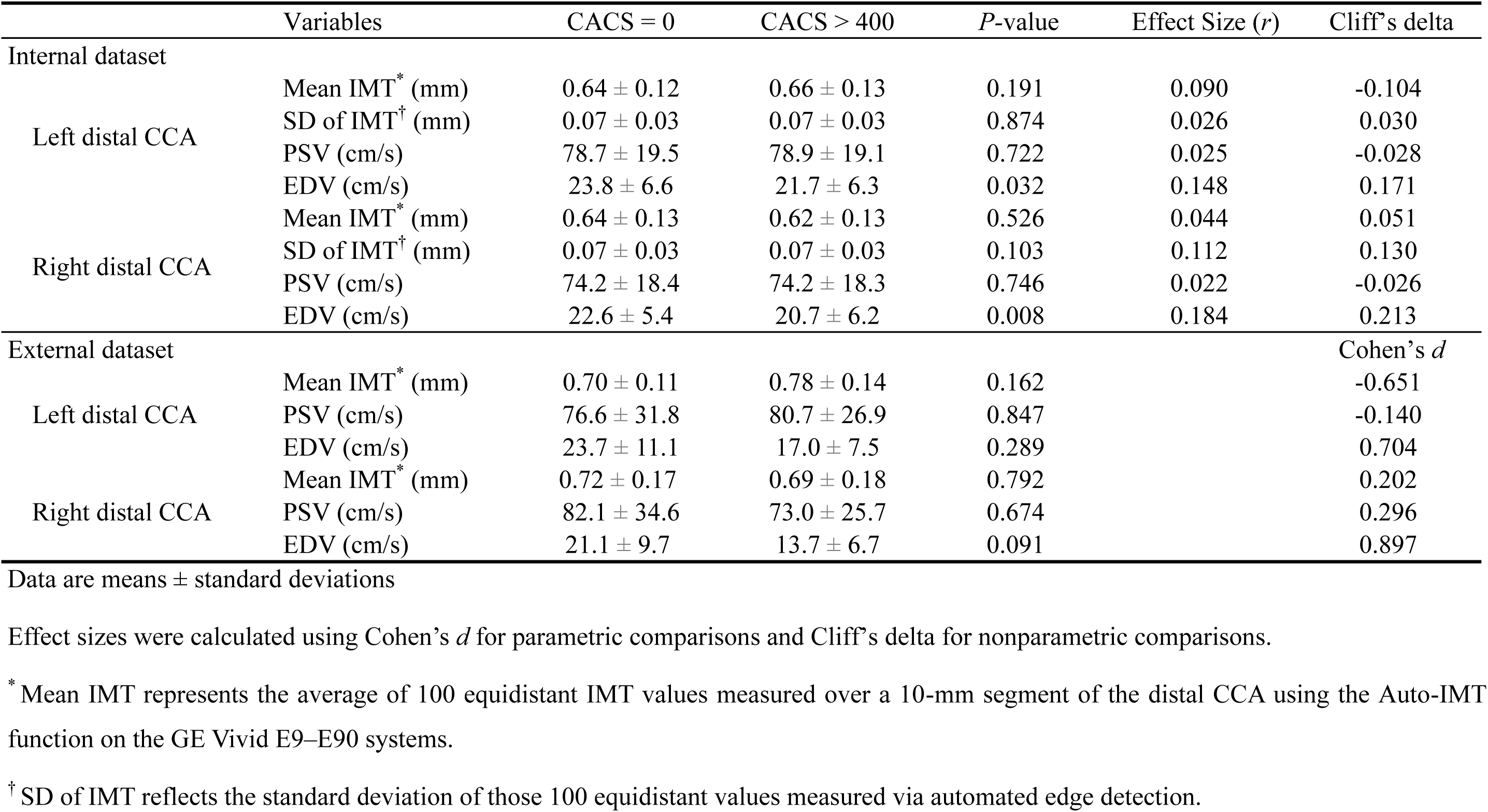
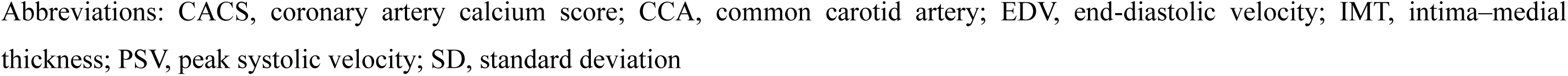
Comparison of conventional CUS measurements between CACS = 0 and CACS > 400 groups in the internal dataset.

### 3.3. Feature selection

A bin width of 0.3 yielded optimal feature extraction results. After removing features with low variance (threshold < 0.2) and high inter-feature correlation (threshold > 0.9), 42 to 46 features remained. Then, final selection using bootstrap-based ranking for reproducibility resulted in 7 to 11 stable features per group (all *p* < 0.05). The number of features retained after each filtering step is shown in **Figure 2**. A complete list of the selected features, statistically significant and ranked by FI, is presented in **Figure 3**. The overall radiomics pipeline, including segmentation, preprocessing, filtering, feature extraction, and feature selection, is summarized in **Graphic Abstract B**.

**Figure 2.**
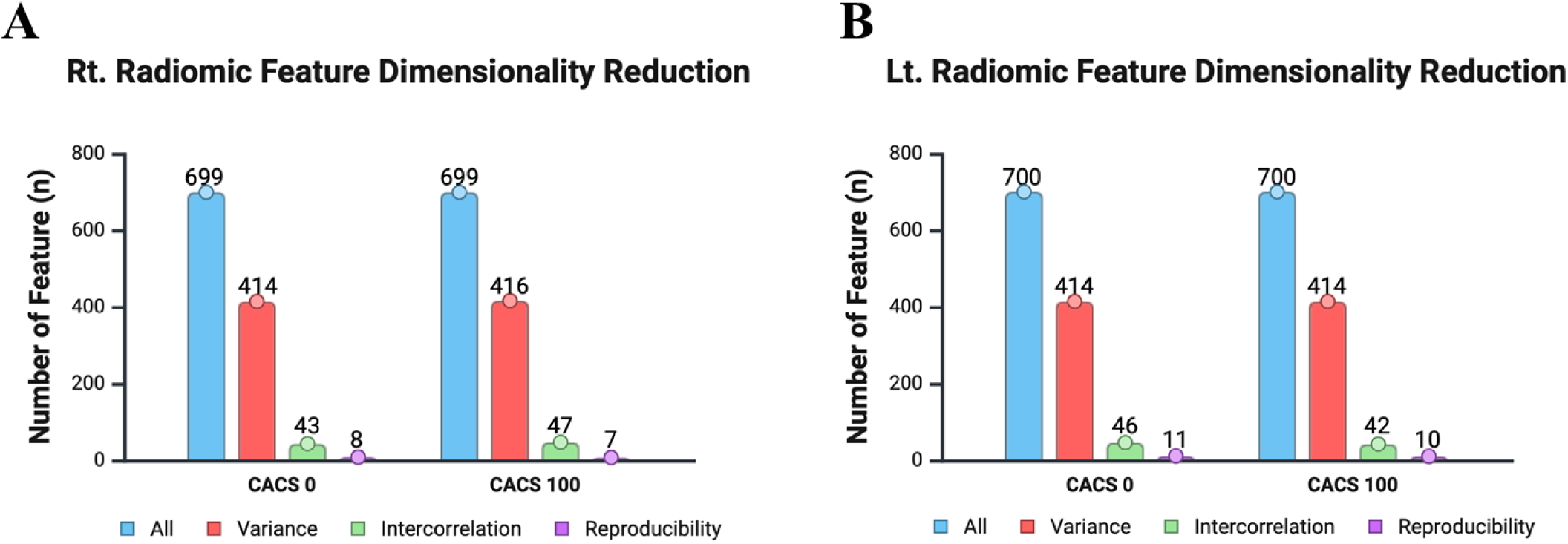
Stepwise dimensionality reduction of radiomic features from the right (A) and left (B) CCA ultrasound images. From an initial set of 700 features, sequential filtering based on low variance, high inter-feature correlation, and reproducibility resulted in 7–11 stable features per group. Abbreviation: CCA, common carotid artery

**Figure 3.**
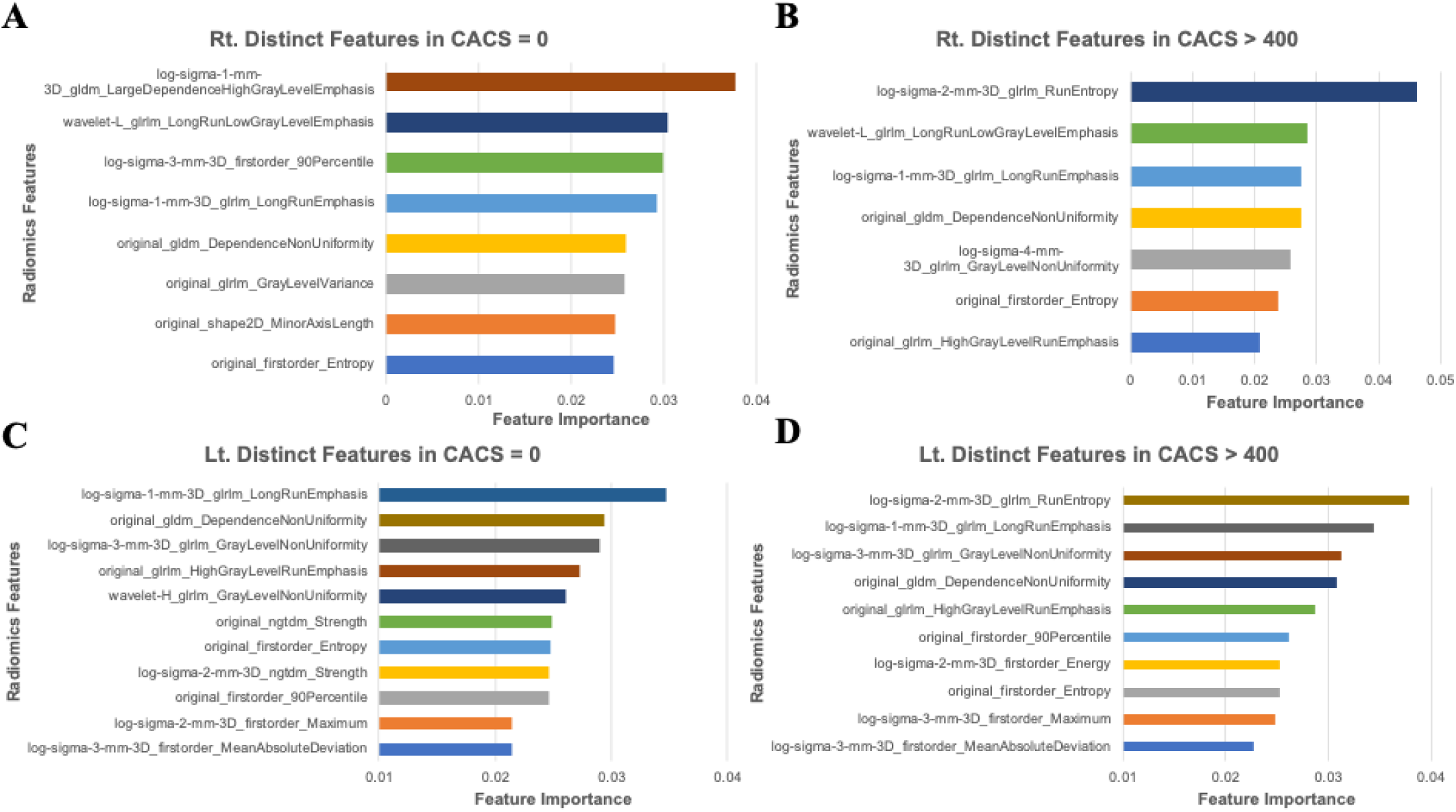
Top-ranked radiomic features distinguishing CACS = 0 and CACS > 400 groups in the right (top) and left (bottom) distal CCA. Feature importance values were derived from bootstrap-based ranking. (A), right Distinct features in CACS = 0 group; (B), right Distinct features in CACS > 400 group; (C), left distinct features in CACS = 0 group; (D), left distinct features in CACS > 400 group. Abbreviation: CACS, coronary artery calcium score; CCA, common carotid artery

#### 3.3.1. Group-wise differences in common radiomic features

Radiomic features commonly identified in both CACS = 0 and CACS > 400 groups (**Figure 4A, B)** were further evaluated for group-wise differences within each carotid artery. In the right distal CCA, all four commonly selected features-originally identified in both groups-exhibited statistically significant differences between CACS = 0 and CACS > 400 (all *p* < 0.05 to < 0.01; **Figure 5A-D**). Similarly, in the left distal CCA, all seven common features demonstrated significant intergroup differences (all *p* < 0.05 to < 0.01; **Figure 6A-G**).

**Figure 4.**
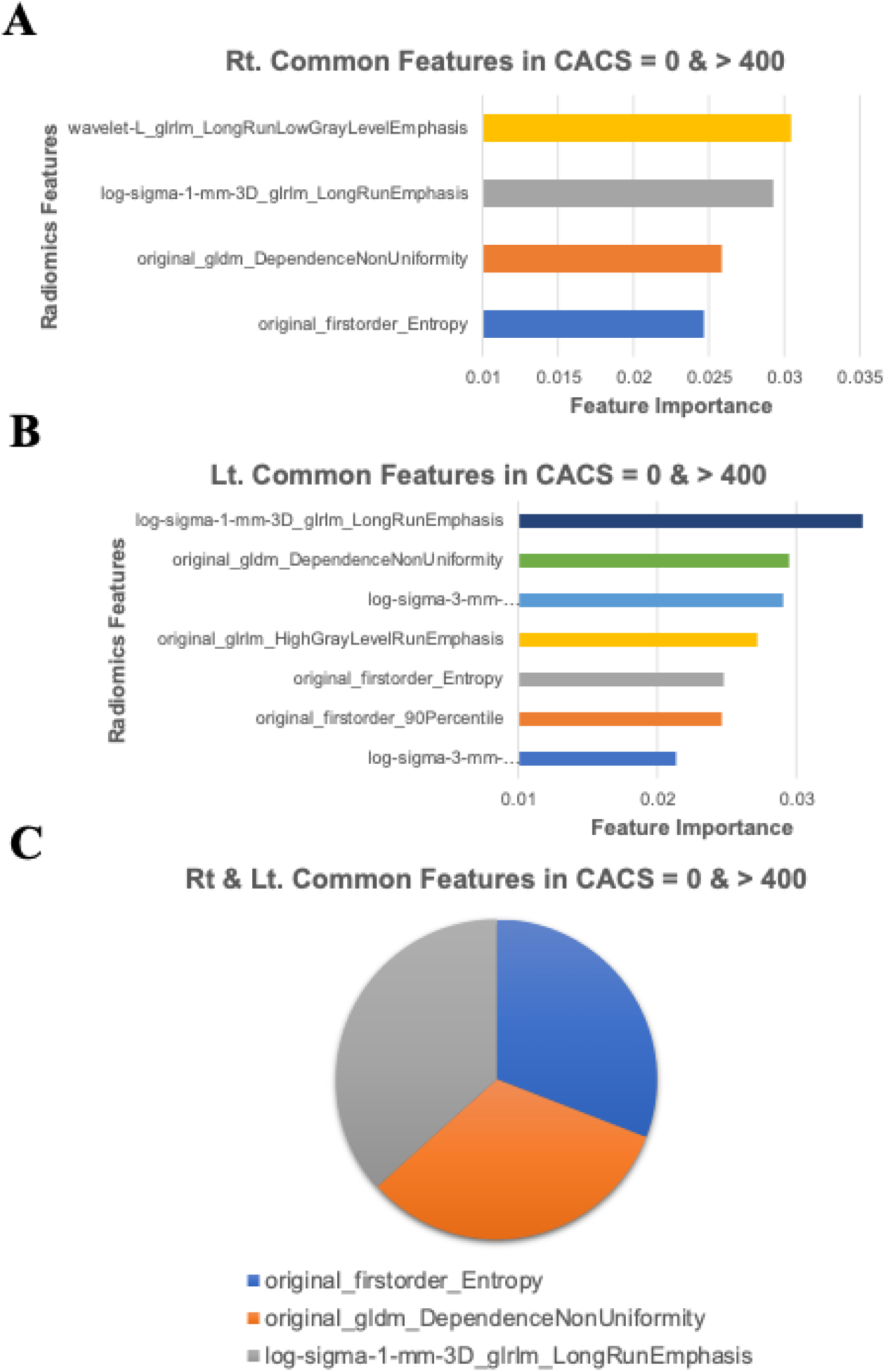
Radiomic features consistently identified in both CACS = 0 and CACS > 400 groups within the right (A) and left (B) CCAs, as well as common features across both sides (C). Abbreviation: CACS, coronary artery calcium score; CCA, common carotid artery

**Figure 5.**
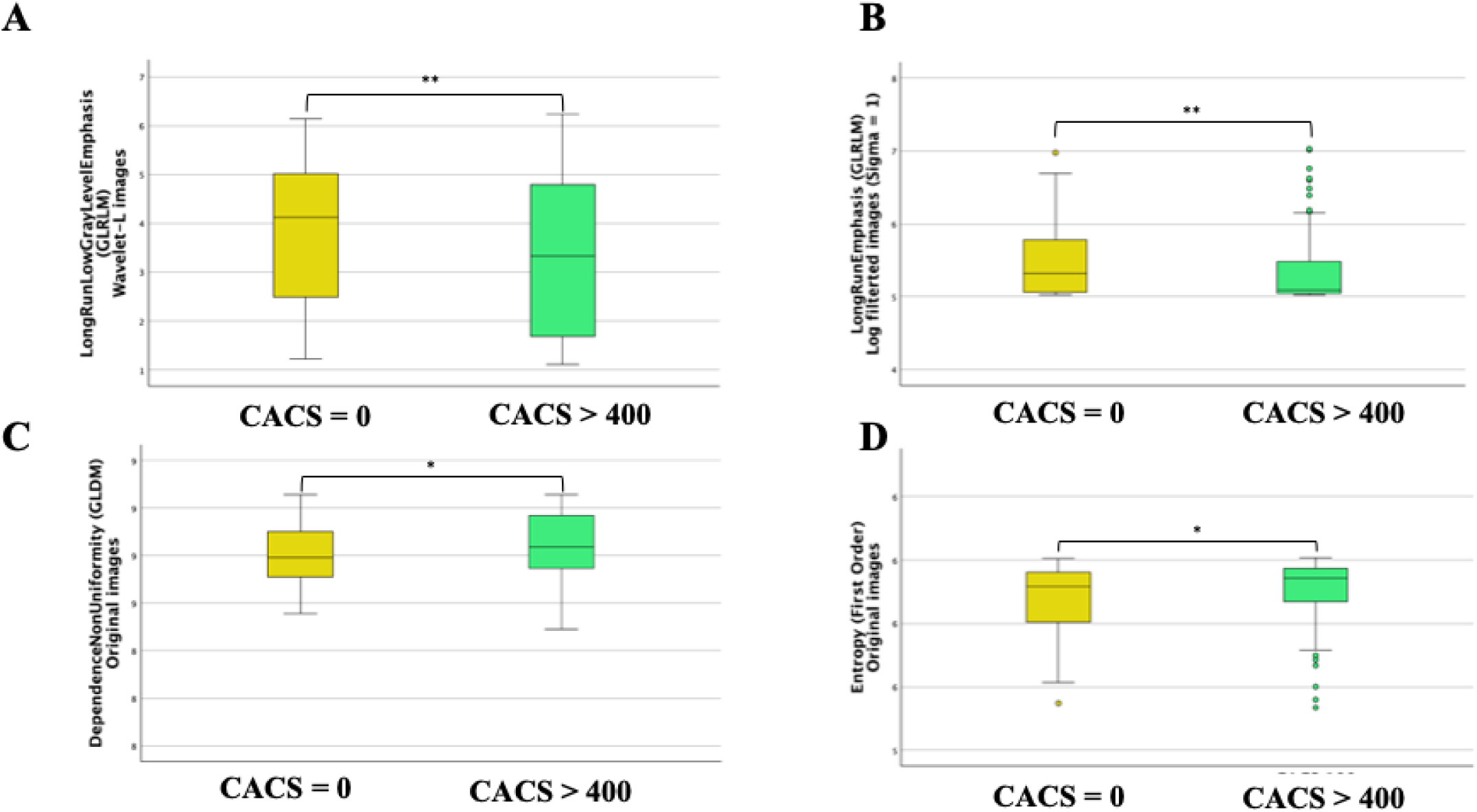
Radiomic features that showed significant differences between CACS = 0 and CACS > 400 groups in the right CCA. (A), Long Run Low Gray Level Emphasis (GLRLM) from wavelet-L images; (B), Long Run Emphasis (GLRLM) from log filtered images (*s* = 1); (C), Dependence Non-Uniformity (GLDM) from original images; (D), Entropy (first-order) from original images. Abbreviation: CACS, coronary artery calcium score; CCA; GLDM, gray-level dependence matrix; GLRLM, gray-level run length matrix; *s*, sigma

**Figure 6.**
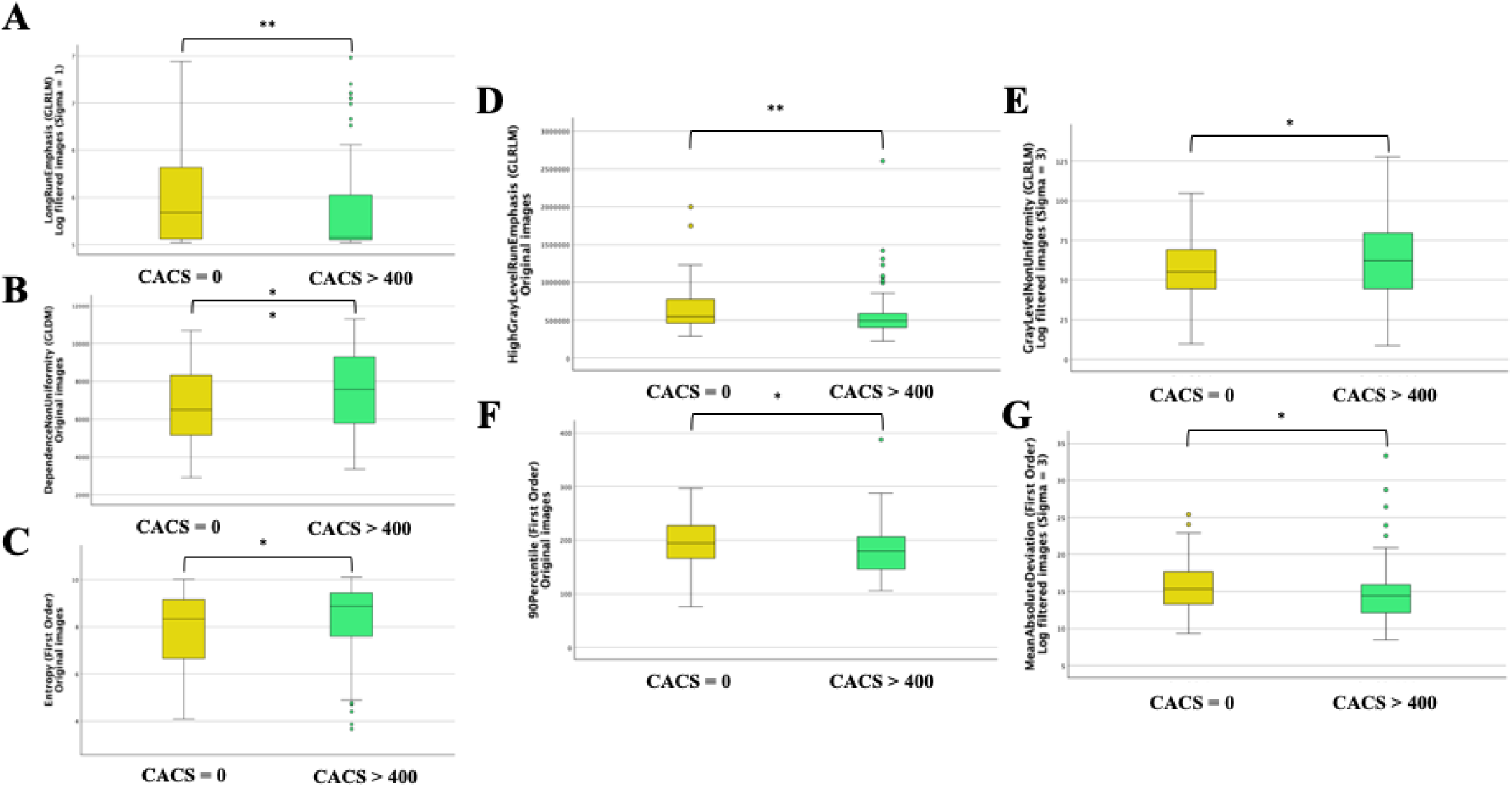
Radiomic features that showed significant differences between CACS = 0 and CACS > 400 groups in the left distal CCA. (A), Long Run Emphasis (GLRLM) from log filtered images (*s* = 1); (B), Dependence Non-Uniformity (GLDM) from original images; (C), Entropy (first order) from original images; (D) High Gray Level Run Emphasis (GLRLM) from original images; (E), Gray Level Non-Uniformity (GLRLM) from log filtered images (*s* = 3); (F), 90th Percentile (first order) from original images; (G), Mean Absolute Deviation (first order) from log filtered images (*s* = 3). Abbreviation: CACS, coronary artery calcium score; CCA; GLDM, gray-level dependence matrix; GLRLM, gray-level run length matrix; *s*, sigma

Figure 4C illustrates the subset of features that were consistently identified in both carotid arteries across both groups. Three features, Entropy (First Order), Dependence Non-Uniformity (GLCM), and Long Run Emphasis (GLRLM, log-filtered, *σ* = 1) (all *p* < 0.05).

#### 3.3.2. Common carotid arteries in the CACS = 0 group

In the CACS = 0 group, four radiomic features had significantly higher values than in the CACS > 400 group (all *p* < 0.05) and were bilaterally consistent: 90th Percentile (First Order), Long Run Emphasis (GLRLM), Dependence Non-Uniformity (GLDM), and Entropy (First Order). These features were selected based on high FI and consistent expression across both sides, indicating potential utility as stable markers of vascular texture symmetry in individuals without CAC (**Table 3, S2)**.

**Table 3.** Side-specific and bilaterally consistent radiomic features in CACS = 0 Group.

| Radiomics Features | Left distal CCA |  |  | Right distal CCA |  |  |
| --- | --- | --- | --- | --- | --- | --- |
|  | FI | Feature value | *P*-value | FI | Feature value | *P*-value |
| 90th Percentile (First order) | 0.025 | 27.022 | 0.021 | 0.030 | 29.256 | 0.007 |
| Long Run Emphasis (GLRLM) | 0.035 | 7.509 | 0.005 | 0.030 | 7.275 | 0.007 |
| Dependence Non-Uniformity (GLDM) | 0.030 | 6695.063 | 0.003 | 0.026 | 6612.502 | 0.015 |
| Entropy (First order) | 0.025 | 7.811 | 0.019 | 0.025 | 7.733 | 0.028 |

**Note.** Feature importance (FI) was calculated using bootstrap-based ranking. Group-wise comparisons were performed using the Mann–Whitney U test. 90th Percentile is derived from log filtered images ( $\sigma = 3$ ), Long Run Emphasis is from log filtered images ( $\sigma = 1$ ), Dependence Non-Uniformity is from original images, and Entropy is from original images.
Abbreviation: CACS, coronary artery calcium score; CCA, common carotid artery; EDV, end-diastolic velocity; FI, feature importance; GLDM, gray-level dependence matrix; GLRLM, gray-level run length matrix; IMT, intima–medial thickness; PSV, peak systolic velocity; $\sigma$ , sigma.

Among them, the 90th Percentile feature (log-filtered, σ = 3) appeared only in the CACS = 0 group and was absent in the CACS > 400 group across both datasets. In the internal dataset, values were higher on the right CCA than the left (29.256 vs. 27.022), whereas in the external dataset, the left CCA exceeded the right (27.02 vs. 24.79), indicating preserved vascular integrity with group-specific indicator.

#### 3.3.3. Common carotid arteries in the CACS > 400 group

In the CACS > 400 group, five radiomic features were identified as statistically significant in the internal dataset (all *p* < 0.05) and reproducibly selected from both distal CCAs, showing consistent trends across internal and external datasets: Run Entropy (GLRLM), Long Run Emphasis (GLRLM), Entropy (First Order), Dependence Uniformity (GLDM), and High Gray Level Run Emphasis (GLRLM) (Table 4). These features may indicate their potential as robust markers of radiomic alterations associated with high CAC burden (**Table 4, S3)**.

**Table 4.** Side-specific and bilaterally consistent radiomic features in CACS > 400 Group.

| Radiomics Features | Left distal CCA |  |  | Right distal CCA |  |  |
| --- | --- | --- | --- | --- | --- | --- |
|  | FI | Feature value | *P*-value | FI | Feature value | *P*-value |
| Run Entropy (GLRLM)) | 0.038 | 8.475 | 0.002 | 0.046 | 8.441 | 0.001 |
| Long Run Emphasis (GLRLM) | 0.034 | 5.565 | 0.005 | 0.028 | 6.382 | 0.007 |
| Entropy (First order) | 0.025 | 8.265 | 0.019 | 0.024 | 8.191 | 0.015 |
| Dependence Non-Uniformity (GLDM) | 0.031 | 7498.088 | 0.003 | 0.028 | 7251.677 | 0.028 |
| High Gray Level Run Emphasis (GLRLM) | 0.029 | 550846.9 | 0.003 | 0.021 | 535747.2 | 0.017 |

**Note.** Feature importance (FI) was calculated using bootstrap-based ranking. Group-wise comparisons were performed using the Mann–Whitney U test. Run Entropy is from log filtered images ( $\sigma = 2$ ), Long Run Emphasis is from log filtered images ( $\sigma = 1$ ), Entropy is from original images, Dependence Non-Uniformity is from original images, and High Gray Level Run Emphasis is from original images.
Abbreviation: CACS, coronary artery calcium score; CCA; FI, feature importance; GLDM, gray-level dependence matrix; GLRLM, gray-level run length matrix

Among the selected features, both Run Entropy (GLRLM, log-filtered, *σ* = 2) and High Gray Level Run Emphasis (GLRLM) were uniquely observed in the CACS > 400 group and not selected in the CACS = 0 group from both datasets. Run Entropy was higher in the external dataset compared to the internal dataset for the right CCA (9.530 vs. 8.441) but slightly lower for the left CCA (8.616 vs. 8.475). High Gray Level Run Emphasis was elevated in the external dataset for the right CCA (588,747.7 vs. 535,747.2) but reduced for the left CCA (543,060.7 vs. 550,846.9), reflecting group-specific markers while maintaining the overall pattern of intensified high-gray-level textures in individuals with severe calcification.

Notably, Dependence Non-Uniformity (left) exhibited statistically significant differences between CACS = 0 and CACS > 400 groups in both the internal (both, *p* = 0.003) and external (CACS = 0, *p* = 0.048; CACS > 400, *p* =0.045) datasets, reinforcing its potential as a robust and generalizable radiomic feature (**Table 3, 4, S2, S3).** Based on closely matched conventional CUS measures (CAC=0, left CCA: mean IMT 0.73 mm, PSV 74.3 cm/s, EDV 24.1 cm/s; right CCA: mean IMT 0.72 mm, PSV 78.7 cm/s, EDV 28.4 cm/s; HbA1c 5.2%, SBP 118 mmHg) and a CAC>400 case with comparable values (left CCA: mean IMT 0.75 mm, PSV 72.2 cm/s, EDV 31.7 cm/s; right CCA: mean IMT 0.73 mm, PSV 77.6 cm/s, EDV 32.8 cm/s; HbA1c 5.6%, SBP 116 mmHg), the images appear similar by conventional ultrasound metrics. Nevertheless, radiomics maps demonstrated distinct texture patterns in the CAC>400 case, supporting that radiomics can discriminate risk even when standard measures are closely matched **(Graphic Abstract C).**

## DISCUSSION

This study demonstrates that radiomics-based analysis of CUS identified subclinical vascular texture differences in patients with high coronary calcium burden who would otherwise be classified as low-risk based on the absence of carotid plaque. Radiomic features incorporating the spatial distribution of voxels, such as Entropy, Dependence Non-Uniformity, and Long Run Emphasis were consistently identified across both common carotid arteries and in both CACS groups, emerging as robust and reproducible texture markers (Entropy: measures disorganized intensity patterns suggestive of tissue heterogeneity; Dependence Non-Uniformity: captures irregular patch sizes within the arterial wall; Long Run Emphasis: reflects elongated uniform structures that may relate to fibrotic or calcified streaks). These features may correspond to irregularities in grayscale intensity distribution and structural uniformity, potentially reflecting early arterial remodeling, fibrous cap disruption, or microcalcification—key features of subclinical atherosclerosis (29). Group-specific features such as 90th Percentile (CACS = 0) and Run Entropy (CACS > 400) suggest the presence of distinct microstructural patterns that may reflect preserved versus heterogenous vascular integrity. Compared to conventional ultrasound metrics (e.g., IMT, EDV), which showed limited or no statistically significant differences between CACS groups in both datasets, radiomic analysis identified multiple features with significant intergroup differences, similarities, and higher discriminatory power between right and left distal CCA. This highlights the added value of radiomics in detecting subclinical vascular changes and supports its potential to enhance cardiovascular risk stratification in asymptomatic individuals.

Previous studies have highlighted a frequent mismatch between CUS and CACS, where elevated coronary calcium may occur despite normal-appearing carotid arteries (15, 16). Despite this gap, few studies have applied radiomics to address it. While radiomics has been increasingly explored in CT and MRI for cardiovascular risk assessment (30–32), its application to ultrasound—particularly in a preventive screening context—remains sparse. Regarding coronary artery disease assessment using CUS radiomics, a recent study demonstrated successful evaluation of coronary disease severity using the SYNTAX score, achieving excellent performance (AUC 0.939) when combining radiomics features with clinical factors (33). This approach identified key texture features reflecting internal plaque characteristics invisible to conventional assessment, focusing on patients with existing carotid plaques rather than subclinical changes in normal-appearing arteries. Our study contributes to the growing body of evidence by demonstrating that CUS radiomics captures vascular texture alterations not visible through conventional imaging, even in plaque-free arteries, offering a non-invasive, accessible alternative for improving subclinical risk detection.

Several group-specific radiomic features may reflect distinct vascular phenotypes related to CAC burden. In the CACS > 400 group, features such as Run Entropy (textural irregularity) and High Gray Level Run Emphasis (presence of intense, clustered signals) suggest increased vascular heterogeneity, which could correspond to early signs of vascular remodeling or microcalcification not evident in conventional imaging. Conversely, 90th Percentile (upper-range signal intensity), observed only in the CACS = 0 group, may represent preserved vascular integrity and homogeneous tissue texture. These findings support the potential of radiomic texture features as surrogate markers for subclinical atherosclerotic processes, providing insight into disease progression beyond plaque presence alone.

By analyzing the selected radiomics features in both left and right distal CCAs, the approach accounts for anatomical variability and ensures bilateral consistency of feature selection. Bootstrap-based reproducibility ranking and feature importance analysis were applied to enhance the clinical applicability and interpretability of the identified features (26, 34). Key radiomic patterns were consistently observed in both datasets further reinforces the generalizability of our findings, supporting the broader utility of radiomics-based ultrasound in diverse clinical settings.

Several limitations should be acknowledged. First, due to restricted access to clinical variables in the external dataset, we were unable to perform comparative cross-analyses of ultrasound-derived outcomes, such as those conducted using the internal dataset’s metabolic profiles, which may have further validated the associations. In the external dataset, most of the selected radiomic features that were statistically significant in the internal population did not show significant differences between the CACS = 0 and CACS > 400 groups, likely due to the limited sample size and incomplete clinical data in the external dataset. Dependence Non-Uniformity remained statistically significant in the left distal CCA even in the external dataset, supporting its potential robustness and generalizability. Second, this study employed a cross-sectional design, which inherently limits conclusions to association rather than causation or prediction. Longitudinal studies are warranted to assess whether radiomic features can prospectively predict cardiovascular events or progression of coronary calcification. Third, although several texture features were statistically significant and reproducible, their biological interpretation remains speculative. The underlying histopathological correlates—such as fibrotic changes, early calcification, or lipid infiltration—have yet to be validated, requiring validation through histological or molecular imaging studies (35).

CUS radiomics represents a promising approach to detect subclinical vascular changes associated with severe coronary atherosclerosis, even without visible carotid plaque. Identifying reproducible texture features beyond conventional interpretation may improve cardiovascular risk stratification in asymptomatic individuals.

---

**Figure S1.**
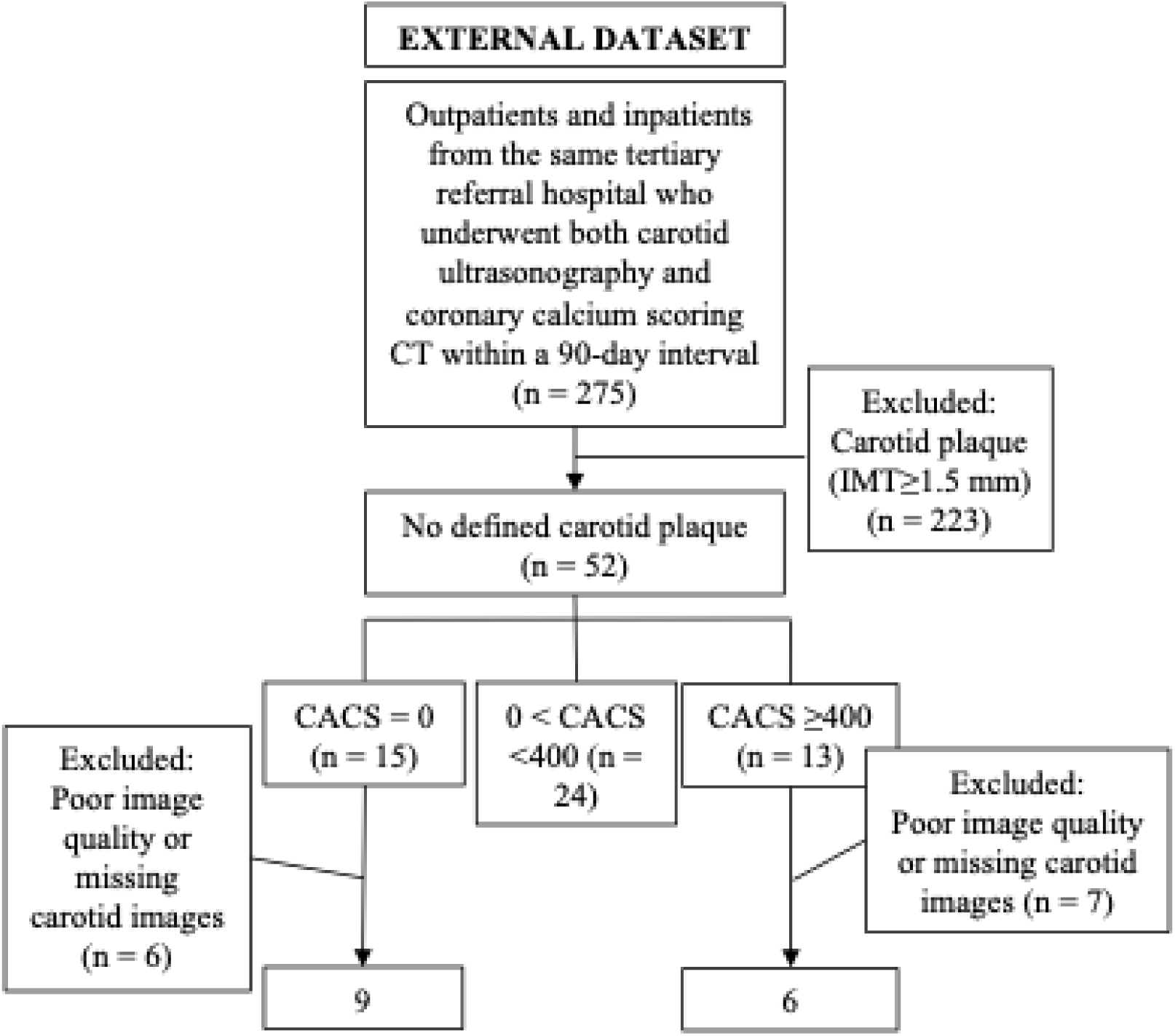
Flow diagrams of the study population selection. External dataset: Inpatients and outpatients (n = 275) from the same tertiary referral hospital (2018–2021) who underwent both tests within a 90-day interval. After applying the same exclusion criteria, 9 participants with CACS = 0 and 6 with CACS > 400 were included. Abbreviation: CACS, coronary artery calcium score; CUS, carotid ultrasound; CVD, cardiovascular diseas

**Figure S2.**
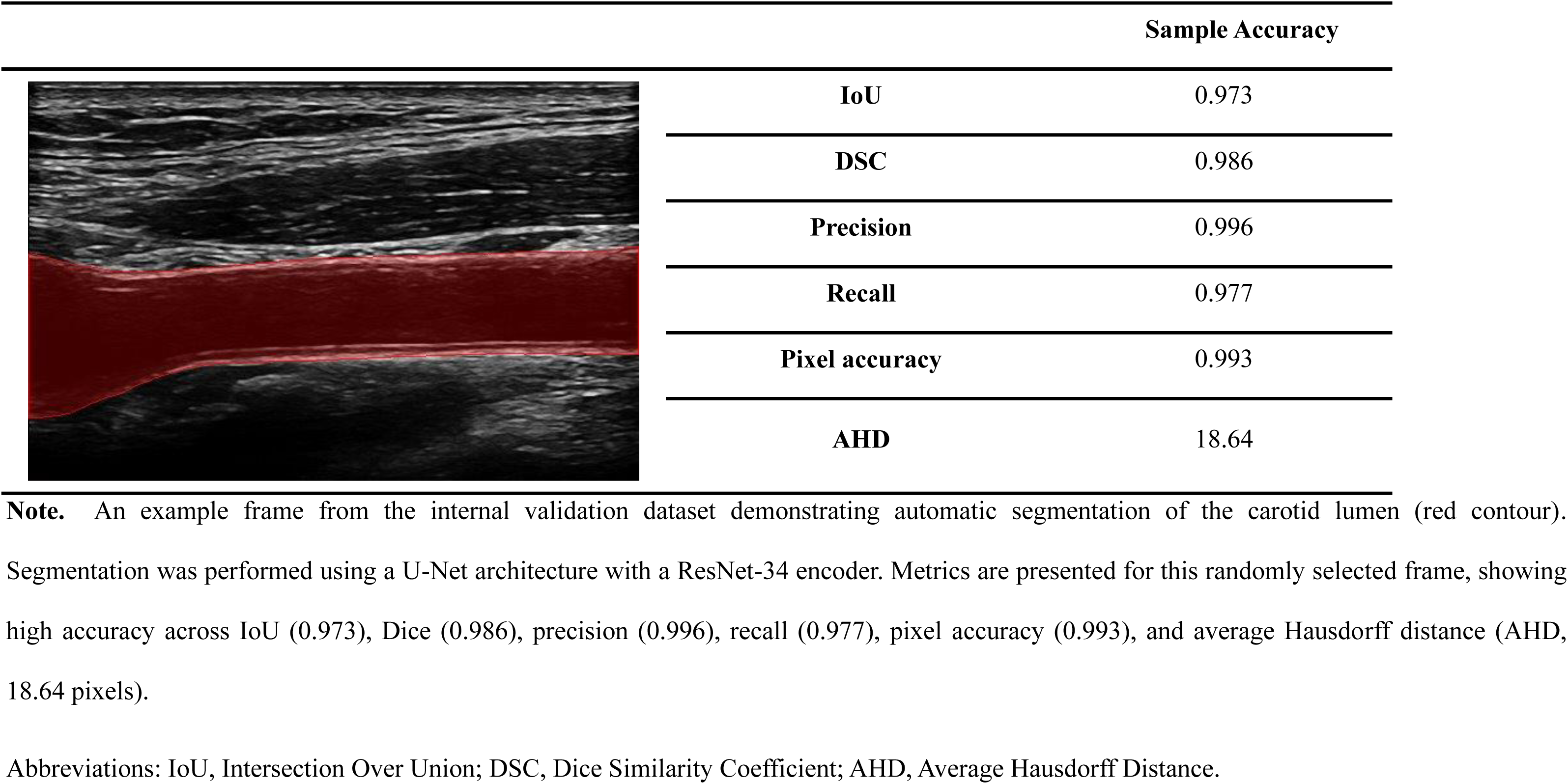
Representative carotid lumen segmentation and overlay result from the internal validation dataset.

